# Time of quickening is associated with the placental site and BMI in nulliparous women

**DOI:** 10.1101/2020.11.27.20239665

**Authors:** Negin Jaafar, Lars Henning Pedersen, Olav Bjørn Petersen, Lone Hvidman

## Abstract

**Introduction:** Quickening, the first sensation of fetal movements, is an important milestone for pregnant women. Information on the expected gestational age at quickening may reduce anxiety and prevent delayed detection of intrauterine demise but the available data are from the 1980s before the emergence of modern ultrasound techniques.

**Materials and methods:** Prospective observational study on nulliparous women blinded for placental location in two hospitals in Denmark. The pregnant women were enrolled at the time of nuchal translucency scan, placental location was determined at time of second trimester scanning.. The women were blinded to placenta location before time of quickening. Time of quickening were reported by 122 women, 65 with an anterior and 57 with a posterior placenta. Thirteen women had a BMI >30 (10.7%).

**Results:** The mean gestational age for quickening was 19 + 0 weeks for nulliparous women. The timing depended on placental site; women with an anterior placenta experienced quickening 6.4 days later than the women with a posterior placenta. BMI > 30 was associated with a later time of quickening.

**Conclusions:** Anterior placental location is associated with delay in experience of fetal movements of 6.4 days and this may further be delayed in women with a BMI>30.

## Introduction

Quickening, the first sensation of fetal movements, is an important milestone for pregnant women, partly because it makes the pregnancy feel “real” for the woman. The milestone is particularly important for nulliparous women with no experience from previous pregnancies. Information on the expected gestational age at quickening may reduce anxiety and prevent delayed detection of intrauterine demise.

From a more theoretical perspective, we already inform women of the potential importance of placental location and BMI for quickening and such information should ideally be quantified and based on more than clinical experience.

There are few published studies on quickening [1-3]. Based on both clinical experience and available evidence, quickening seems to be associated with parity, placental location, and amniotic fluid volume. The expected time and variation in the timing of quickening and, importantly, the magnitude of change because of individual factors remain unresolved. Further, no large prospective studies have been published since 1984 and this renders existing estimates uncertain due to the profound changes in ultrasound techniques.

The aims with this prospective study were to investigate time of quickening in pregnancy and to estimate the potential influence of placental site and maternal BMI in nulliparous woman blinded for placental location.

## Material and Methods

### Cohort

Nulliparous pregnant women were enrolled at the time of nuchal translucency scan (NT scan) at two Danish hospitals (Aarhus University Hospital and Viborg Regional Hospital, Central Denmark Region) from August 2010 to October 2014. All women underwent both NT scan and second trimester malformation scan as part of routine prenatal care. Health care is free in Denmark, and the study had consequently no influence on the availability or quality of the prenatal care received by the women.

The pregnant women were informed about the project verbally by certified sonographers at time of NT scan, and signed a consent form that included written information on the project.

Only nulliparous women with a normal nuchal scan were included. No women were included prior to gestational week 11 + 6 days and the predefined cut off for participation was gestational week 15 + 0.

All women were scanned according to standard guideline for NT scans in Denmark, except that the pregnant women were blinded for the location of the placenta. Care was taken to avoid accidentally revealing the placental location, verbally or on the visible live scanning. Location of the placentas used in the analyses was determined at the second trimester scan. However, women with potential placenta previa and abnormal pregnancy were informed and excluded. Women with fundal and lateral placentas were excluded from the analyses.

### Quickening

Quickening was defined as the day when the pregnant woman felt fetal movements for first time. The women recorded the timing prospectively and gave the information to the sonographer at time of the second trimester scanning. If no movements had been felt by the time of the second trimester ultrasound scan, the pregnant women were asked to provide the date of quickening by e-mail. Women that provided no information per e-mail were excluded. We included only information on quickening from the women that were certain about the timing; if a woman was unsure, the timing was coded as missing.

### Covariates

Information on maternal age, BMI, occupation, and smoking status was obtained as a routine part of the risk calculation after the NT scan. The estimated day of delivery (EDD) was based on the crown-rump length from the NT scan for all pregnancies. The beginning of gestation was estimated by subtraction 280 days from the EDD.

### Statistics

Difference in timing of quickening by placental location (anterior or posterior) or BMI (≤30 or >30) was compared using non-parametric Ranksum test (Mann-Whitney). The risk of cesarean section depending on placental location was compared using Fisher’s exact test. We used linear regression models to estimate the difference in timing adjusted for potential confounders. The Kaplan-Maier estimator was used to investigate timing stratified by placental location.

### Ethics

The Danish data protection agency and the Regional Scientific Ethical Committee (Aarhus, Denmark) approved the study (J. nr.2012-41-1357).

## Results

We included 136 nulliparous women with a normal nuchal scan. Fourteen were excluded (10.3%): nine women provided no information on quickening, three were excluded because of placenta praevia, and two due to uncertain date of quickening. Of the 122 women included 65 had an anterior and 57 had a posterior placenta.

The mean gestational age at quickening for nulliparous women was 133 days (ranges from 96-175 days, interquartile ranges from126-141 days). Stratified by the placental location, mean gestational age at quickening among women with an anterior placental location was 136.1 days (interquartile ranges from 128-144 days) and 129.7 days (interquartile ranges from 125-136 days) among women with a posterior placenta (Figure 1). The difference was statistically significant (p=0.005).

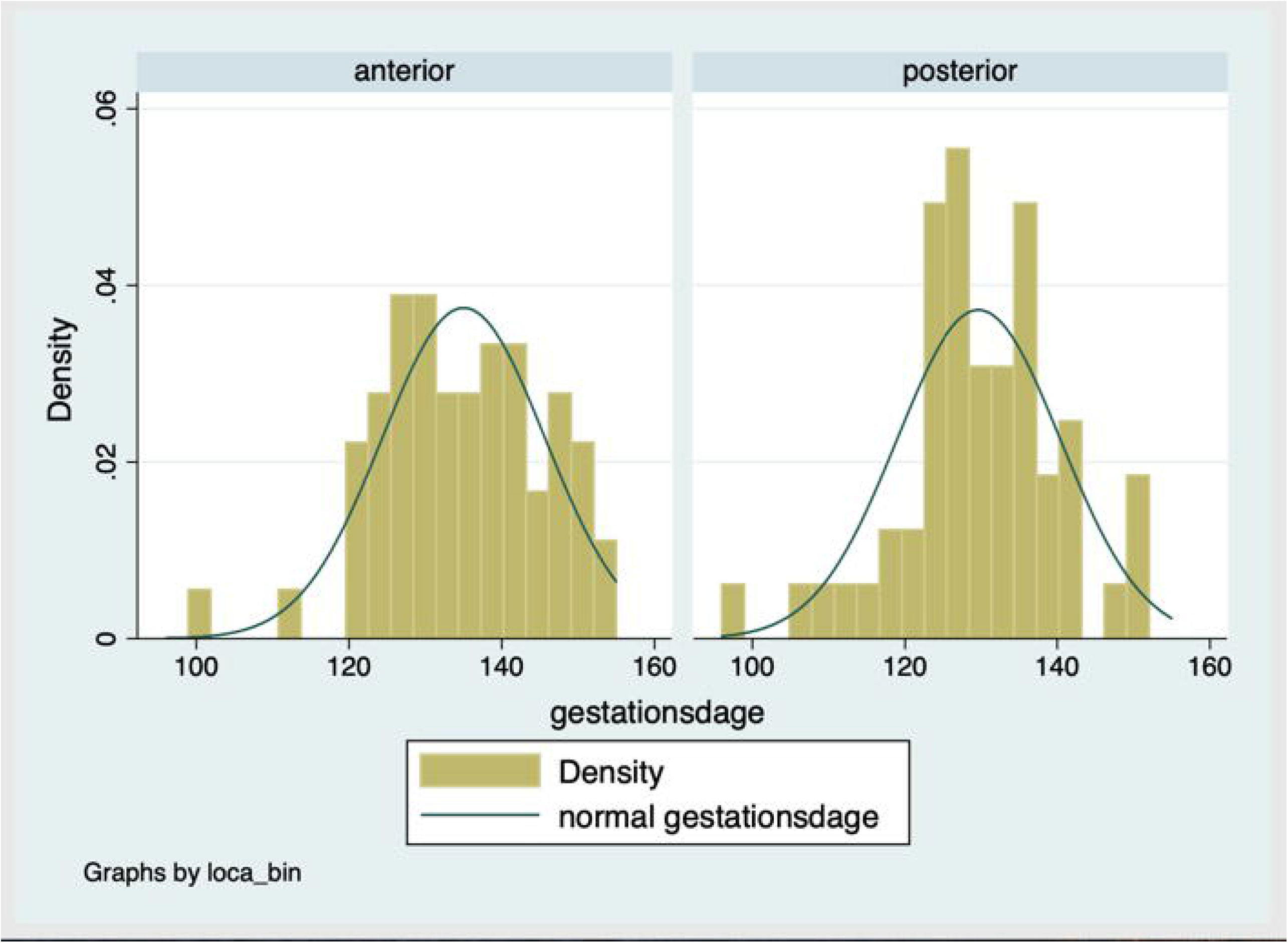

Women with BMI ≥30 experienced quickening 6.0 days later than women with BMI<30 (p<0.05). The difference was statistically significant in a linear regression model including BMI and the placental location as independent dichotomised variables (BMI coef. 7.5 days, 95% confidence interval 0.5 −14 days). Figure 2 illustrates the association between BMI and timing of quickening. The lowess regression suggest a potential association with BMI>30 but the coefficients for BMI <30 and >=30, respectively, were not statistically significant.

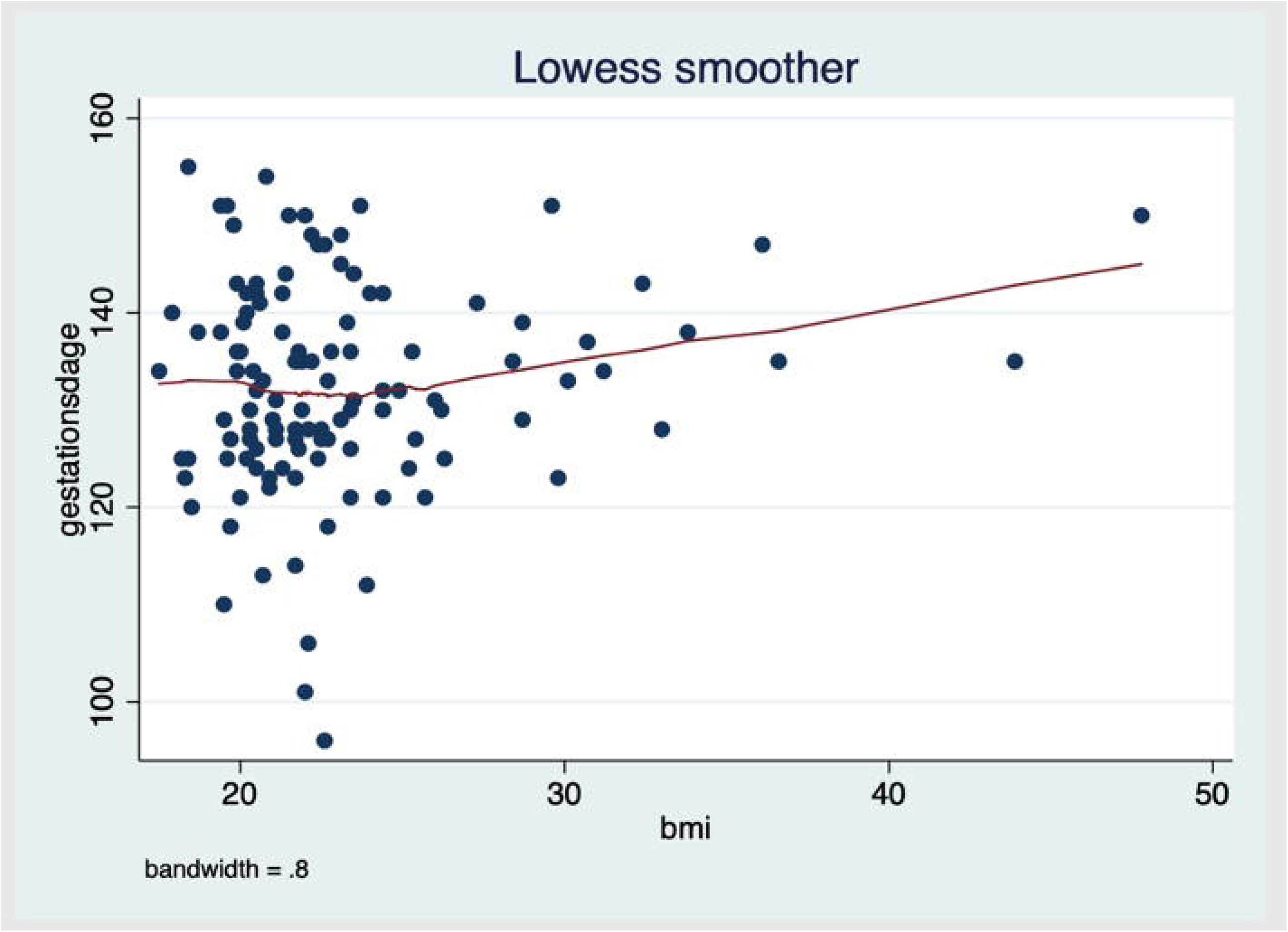

There was a statistically significant different risk of cesarean section dependent on location of the placenta among the women with information of mode of delivery. The proportion of women that had a cesarean section was 8.6% and 26.5% among women with an anterior and posterior placenta, respectively (p=0.013). We had no prior hypothesis on the potential association between placental location and risk of caesarean section and the result may be a chance finding.

## Discussion

Women with an anterior placenta felt the first fetal movements 6.4 days later than those with a posterior placenta. BMI > 30 was associated with quickening 6.0 days later in pregnancy but sample size was limited.

The results suggest that an anterior placenta delays the maternal detection of fetal movements. This might be explained by the fact that the anterior wall of the uterus has more room for distention in the abdomen than does the posterior wall or that quickening at this stage partly relies on sensation in the abdominal wall. We speculate that the association suggested for BMI>30 is due to intra-abdominal fat reflecting the same mechanism. Previous studies are in accordance with this.

In 1979, Hertogs et al [2] evaluated the correlation between maternal perception of fetal movement and live ultrasound imaging among 20 women (nulliparous and multiparous) between 32 and 43 weeks of gestation. Nineteen of the 20 women had obstetric complications and only one woman had normal pregnancy. They found that women with an anterior placental site did perceived 55% of fetal movements, whereas those with a posterior site perceived 68%. This was not statistically significant (p< 0.3) potentially due to the small sample size [2].

Neldam et al [3], in a study published in 1984, compared fetal movements detected by ultrasound with those felt by the pregnant women in gestational week 20 to 23. They found differences between maternal experience and ultrasound estimation only in pregnancies with an anterior placenta but in pregnancies with a posterior placenta there was no differences [3].

In 1984, Gillieson et al [1] studied 112 women, including 51 nulliparous, evaluating the placental site and parity influences the date of quickening. They found that quickening occurred approx. one week earlier among women with a posterior placenta. The gestational age in that study is expected to be less precisely calculated compared to the estimates based on modern ultrasound techniques used in our study but, taken that into account, the similarity is striking [1].

There are few data on the potential significance of maternal body composition. Tuffnel et al [4] found that women weighing over 80 kg were more likely to report decreased fetal movements, corroborating that increasing maternal weight may be associated with a decreased sensitivity of movement perception rather than actual decrease in movement. They speculate that the sensation of fetal movements arises from pressure against body wall structures rather than the uterus[4].

We found an association between placental location and risk of cesarean section. The analysis was planned prior to the collection of the data but we had not prior hypothesis and no mechanistic explanation for the result. It may well be a change finding and needs to be corroborated in larger datasets.

## Limitations

Nine women reported no information on quickening (6.8%), five woman with anterior placental site, and 4 woman with posterior placental site, and this might lead to bias if the loss to follow-up was associated with placenta site and time of quickening. Reassuringly, because of the blinding of the women such bias seems unlikely and, importantly, the size of the loss to follow-up is small.

Sample size may limit the power of the study and e.g. lack of association with BMI <30 might be due to type 2 error. Residual confounding is always problematic in observational studies but such potential confounders have to be associated with placental site. We know of no factors in nulliparous pregnancies that both explain the position of the placenta and is associated with quickening. In studies including multiparous women there is a theoretical association between previous cesarean section and the propensity for specific placental location, however, to our knowledge, previous cesarean section has not been associated with time of quickening We further found equal distribution of anterior and posterior placental site (in accordance with e.g. Hohler et al [5]) which is reassuring.

## Conclusion

The mean gestational age for quickening was 19 + 0 weeks for nulliparous women. The timing depended on placental site; women with an anterior placenta experienced quickening 6.4 days later than the women with a posterior placenta. BMI > 30 was associated with a later time of quickening. In our study, placental position in nulliparous women were associated with the risk of cesarean section, a result that needs to be replicated in a different study population.

## Data Availability

Anonomized data is available upon request. First author is responsible for distribution of such data.

